# VAL-1221 FOR THE TREATMENT OF PATIENTS WITH LAFORA DISEASE: STUDY PROTOCOL FOR A SINGLE-ARM, OPEN-LABEL CLINICAL TRIAL

**DOI:** 10.1101/2024.02.01.24302141

**Authors:** Lorenzo Muccioli, Luca Vignatelli, Maria Tappatà, Serena Mazzone, Corrado Zenesini, DEFEAT-LD study group, Dustin Armstrong, Roberto Michelucci, Francesca Bisulli

**Author notes:** Corresponding Author: Francesca Bisulli, MD, PhD Address: Ospedale Bellaria, Via Altura 3, 40139 Bologna.

## Abstract

**Introduction:** Lafora Disease (LD) is an ultrarare fatal progressive myoclonic epilepsy, causing drug-resistant epilepsy, myoclonus, and psychomotor deterioration. LD is caused by mutations in EPM2A or NHLRC1, which lead to the accumulation of polyglucosans in the brain and neurodegeneration. There are no approved treatments for LD. VAL-1221 is a fusion protein comprised of the Fab portion of a cell-penetrating antibody and recombinant human acid alpha glucosidase, and has demonstrated an ability to clear polyglucosans. We hypothesize that intravenous infusion of VAL-1221 might be able to degrade cerebral polyglucosans and stabilize or improve disease outcomes. The aim of this study is to assess the safety and preliminary efficacy of VAL-1221 in patients with LD.

**Methods and analysis:** The study is a phase 2, single-arm, open-label, baseline-controlled clinical trial which will be conducted in a single investigational study center in Italy, namely the sponsor “IRCCS Istituto delle Scienze Neurologiche di Bologna - Azienda USL di Bologna”. The study will enroll 6 genetically-confirmed patients with mid- to late stage LD. The global duration of the study for each participant will be 18 months, including screening period, open-label treatment (12 months), and follow-up period. VAL-1221 20 mg/kg will be administered as an intravenous infusion every week for 3 weeks, then every other week. Patients will undergo full clinical assessments at baseline, at an intermediate and at the end-of-treatment visit. The primary objective is to evaluate the safety. The exploratory efficacy endpoints will be related to epilepsy, neuropsychological and motor functions, global assessment and disease burden, in addition to biomarkers. Statistical analyses will be primarily descriptive.

**Ethics and dissemination:** The study protocol was approved by the local ethics committee (number 232-2023-FARM-AUSLBO–23020, 22-Mar-2023). The results of this study will be disseminated by the investigators through presentations at international scientific conferences and reported in peer-reviewed scientific journals.

**Trial Registration:** European Union Clinical Trials Register (EudraCT number 2023-000185-34).

**ARTICLE SUMMARY:** *Strengths And Limitations Of This Study:*

- This was the first approved study protocol developed for a potential disease-modifying drug in Lafora disease
- The study will be open label due to the serious and life-threatening nature of the disease and its rarity
- Given the lack of a control group in this study, the exploratory endpoints observed will be baseline-controlled and compared with natural history studies
- The rationale for VAL-1221 dose and schedule comes from experience in Pompe disease clinical trials and expanded access programs

## INTRODUCTION

Lafora Disease (LD) is a rare autosomal recessive form of progressive myoclonus epilepsy for which no curative therapies exist. LD affects previously healthy children or adolescents, causing drug-resistant epilepsy, myoclonus, and psychomotor deterioration, leading to loss of autonomy and death after a median of 6 and 11 years, respectively (Pondrelli et al., 2021). LD is ultra-rare and has prevalence rates highest in Mediterranean countries, notably Italy (Nitschke et al., 2018). According to data provided by the Italian Lafora patient association (AILA), at the time of writing there are 25-30 patients with LD in Italy.

LD is caused by loss of function mutations in *EPM2A*, encoding the glycogen phosphatase laforin, or *NHLRC1*, encoding the E3-ubiquitin ligase malin. The absence of either protein results in poorly branched, hyperphosphorylated glycogen, which by extruding water precipitates, aggregates and accumulates into polyglucosans (Lafora bodies - LBs) in different organs (Nitschke et al., 2018). The accumulation of LBs in the brain drives disease progression, being responsible for neurodegeneration and the clinical manifestations of LD (Ganesh et al., 2002; DePaoli-Roach et al., 2010).

The mean age at disease onset is 13 years (Pondrelli et al., 2021). The first symptoms of LD typically consist in myoclonic, generalized tonic-clonic, and focal visual seizures. Epilepsy gradually worsen and become intractable, leading to episodes of status epilepticus. In addition to classic myoclonic seizures, patients may also develop rest, action and stimulus-sensitive myoclonus, which worsen and becomes nearly continuous at late disease stages. Ataxia typically presents in the first 2-3 years of disease, and together with myoclonus contributes to motor disability. Cognitive decline is invariably present, typically manifesting with difficulties at school, and progressing to profound dementia or a vegetative state. (Turnbull et al., 2016)

Detailed data on natural history of LD are currently unavailable. An international natural history and functional status study of patients with LD (NCT03876522) has followed-up 33 patients over 24 months, which were assessed using standardized, quantitative evaluations to identify useful biomarkers and clinical outcome measures for use in Lafora treatment studies has been completed on April 1, 2022. The results have not been published yet, but there is an intention to do so in the coming months, and investigators have been empowered for final data analysis.

At present there are no approved treatments for LD other than symptomatic therapy and assistive devices. As eliminating LBs rescues the neurological phenotype in LD mouse models [Duran et al., 2014], a therapeutic option is the degradation of accumulated LBs. This goal could be achieved by the delivery of a polyglucan-degrading enzyme to the brain.

VAL-1221 is an investigational medicinal produced by Parasail LLC as a treatment for patients with Pompe disease (glycogen storage disease type II) and LD. VAL-1221 is a fusion protein comprised of the Fab portion of a recombinant humanized cell-penetrating antibody (3E10 IgG) and recombinant human acid alpha glucosidase (rhGAA) (Zhou et al., 2019). rhGAA (Myozyme®/Lumizyme®), which utilizes mannose-6-phosphate receptors (M6PR) to gain entry into lysosomes, is indicated for the treatment of patients with Pompe disease (Llerena et al., 2016). In LD, LBs accumulate primarily in the cytoplasm and clinical manifestations are driven by central neurodegeneration (Nitschke 2018). VAL-1221 is designed to preferentially deliver GAA to both lysosomal and extra-lysosomal compartments in key affected organs. The Fab portion of the antibody was demonstrated to have unique capabilities for cell penetration into the cytoplasm via the ENT-2 receptor, which also facilitates brain endothelial cell penetration and blood-brain-barrier (BBB) transport (Zhou et al., 2019)(Rattray et al., 2021). In LD mouse models, VAL-1221 has demonstrated an ability to clear systemic LBs after intravenous infusion and brain LBs after intracerebroventricular (ICV) administration (under publication, personal communication of the corresponding Author Prof. Matthew Gentry, University of Florida). Unfortunately, the current VAL-1221 formulation is not suitable for human ICV injections. However, VAL-1221 has demonstrated a capability to travel to the brain in detectable levels beyond rhGAA-alone (under publication, personal communication of the corresponding Author Prof. Matthew Gentry, University of Florida) and may be able to degrade brain LBs even if administered intravenously.

Clinical experience with VAL-1221 is limited to a Phase 1/2 study in patients with late-onset Pompe disease [NCT02898753] (Kishnani et al., 2019). This was a three-month randomized, repeat-dose, dose-escalation study with patients receiving VAL-1221 (n=9) with a concurrent Lumizyme control (n=2). The primary outcome of the study was the safety. There were no serious or unexpected adverse events (AEs). The most commonly reported AEs were associated with infusion-related reactions (IRR). Additionally, there are about ten patients with LD globally treated with VAL-1221 via expanded access programs or compassionate use (NCT05930223), in whom the treatment was safe and well tolerated (*unpublished data* and *personal communications*).

We hypothesize that VAL-1221, by degrading cerebral LBs, will be able to stabilize or improve seizure control, cognition, motor outcomes, and disease burden. The aim of this study is to assess the safety and preliminary efficacy of VAL-1221 in patients with LD.

## METHODS AND ANALYSIS

### Registration and guidelines

The study protocol reported here (VAL1221-ITLAFORA-01 version 2.0, April 3, 2023) was written in compliance with the Standard Protocol Items: Recommendations for Interventional Trials (SPIRIT) (Chan et al., 2013) and reports the items required by the World Health Organization Trial Registration Data Set (Appendix). The sponsor (no-profit) is IRCCS Istituto delle Scienze Neurologiche – Azienda USL di Bologna, Bologna, Italy. The trial is registered on the European Union Clinical Trials Register (EudraCT number 2023-000185-34).

### Trial objectives

The primary objective of this study is to assess the safety of VAL-1221 on patients with genetically confirmed LD, in mid-late disease stages during 12 months of treatment. Moreover, exploratory efficacy endpoints of VAL-1221 will be assessed, in terms of changes in seizure frequency and epileptiform discharges, stabilization or improvement of cognitive, motor and neuropsychiatric outcomes, as well as neuroradiological and laboratory biomarkers.

### Trial design

The study is a Phase 2, single-arm, open-label, baseline-controlled clinical trial which will be conducted in a single investigational study center in Italy, namely the sponsor “IRCCS Istituto delle Scienze Neurologiche di Bologna - Azienda USL di Bologna”.

The global duration of the study for each patient will be 80 weeks (approximately 18 months), including screening period (4 ± 3 weeks), open-label treatment (51 weeks, approximately 12 months), and follow-up period (24 weeks, approximately 6 months). Patients will undergo full clinical assessments at Baseline (week 1, before the first infusion), at an intermediate evaluation (week 25), and at the end-of-treatment visit, one week after the last infusion (week 52). A six-month observation period will commence to observe the effect(s), if any, of stopping treatment (six-month follow-up visit: week 76).

### Study treatment

VAL-1221 20 mg/kg will be administered as an intravenous infusion every week for 3 weeks, then every other week until week 51. The infusion will start at an initial rate of 1 mg/kg/h, that will be increased by 2 mg/kg/h every half hour as tolerated to a maximum of 7 mg/kg/h until the desired dose is reached. However, both the ultimate dose and infusion rate may be adjusted to half the original rate or lower, according to standard-of-care and at the discretion of the investigator. VAL-1221 is supplied as a sterile, frozen liquid formulation containing 10.0 mg/mL of VAL-1221. The drug product vials are for single dose use of up to 20 mg and 80 mg of VAL-1221, respectively.

A premedication regimen will be administered 30-60 minutes prior to infusion for the first three doses of the study. The premedication regimen will consist of the following: an H1 antihistamine (cetirizine) and an anti-pyretic medication (paracetamol).

Patients who experience infusion-related reactions may continue on VAL-1221 at a lower dose and may be administered glucocorticoids, antihistamines, antipyretics and anti-emetics, as needed. The rate of infusion may be adjusted to half of the original rate or lower, to reduce the symptoms of the reaction.

The Investigator will record the time and dose of all administrations of VAL-1221.

The Pharmacy will maintain accurate records of all investigational product supplies.

Except as indicated in the eligibility criteria, there are no limitations on concomitant therapies before and/or during the study

### Participants

The study will enroll 6 male or female patients with mid- to late stage LD due to the impossibility of detecting potential treatment effects in patients at early disease stage and at end-stage disease. The stage of disease progression may be evaluated with a disability scale based on the residual motor and mental functions, daily living and social abilities, developed by Franceschetti et al. (2006): 1, mild cognitive and motor impairment, preserved daily living activities, and social interaction; 2, moderate mental decline, limitations in motor activities and limited social interaction; 3, severe mental and motor impairment, needing help in walking and regular assistance in daily living activity, and poor social interaction; and 4, patient wheelchair-bound or bedridden, and no significant daily living activities or social interaction.

The sample size for this study is dictated by the rarity of the condition, and the availability of the drug product VAL-1221, and not by any quantitative statistical considerations or formal sample size calculations.

As the onset of LD occurs in childhood, children/minors will be included. Additionally, as cognitive deterioration due to neurodegeneration is a core clinical feature of LD, vulnerable patients with intellectual disability will also be included.

Subjects who are currently cared for at the IRCCS Istituto delle Scienze Neurologiche di Bologna, Italy, or who will be referred to this Institute by other clinicians following the approval of this protocol, will be considered for enrollment and evaluated for inclusion and exclusion criteria. Informed consent will be obtained from the investigator.

IRCCS Istituto delle Scienze Neurologiche di Bologna is a public research institute, a full member of the European Reference Network for Rare and Complex Epilepsies (EpiCARE), highly specialized in the treatment and research of rare epilepsies, including Lafora disease.

### Study inclusion criteria

1. Documented genetic diagnosis of LD based on biallelic pathogenic variants in the *EPM2A* or the *NHLRC1* gene.
2. Between 12 and 28 years of age
3. Mid- or late-stage in the LD evolution (LD progression scale 2-3)(Franceschetti et al., 2006)
4. Able and willing to comply with the study protocol including:

a. Caregiver/trial partner committed to facilitating patient’s involvement in the study who is reliable, competent, and at least 18 years of age.
b. Adequate cognitive capacity to participate in neuropsychological testing adjusted to their level of functioning

### Study exclusion criteria

1. Any known genetic abnormality confounding the phenotype
2. Subjects with any of the following

a. complete absence of speech OR
b. unable to perform any activities of daily living OR
c. completely bedridden
3. Current participation in an interventional or therapeutic study
4. Pregnancy
5. Receiving an investigational drug within 90 days of the Baseline Visit
6. Prior or current treatment with gene or stem cell therapy
7. Any other diseases which may significantly interfere with the assessment of LD
8. Known hypersensitivity or allergy to any of the components of VAL-1221 and excipients.
9. Have any additional conditions which, in the opinion of the Investigator, would make the subject unsafe for inclusion or could interfere with the subject participating in or completing the study.

### Study procedures

The schedule of activities to be performed during the study is provided in Table 1. The following information will be recorded for all potential patients as part of the screening assessments: informed consent, inclusion/exclusion criteria, demography, medical, neurological and epilepsy history, tonic-clonic seizure frequency in the past six months (from clinical records and/or seizure diary), prior and concomitant medications.

**TABLE 1:**
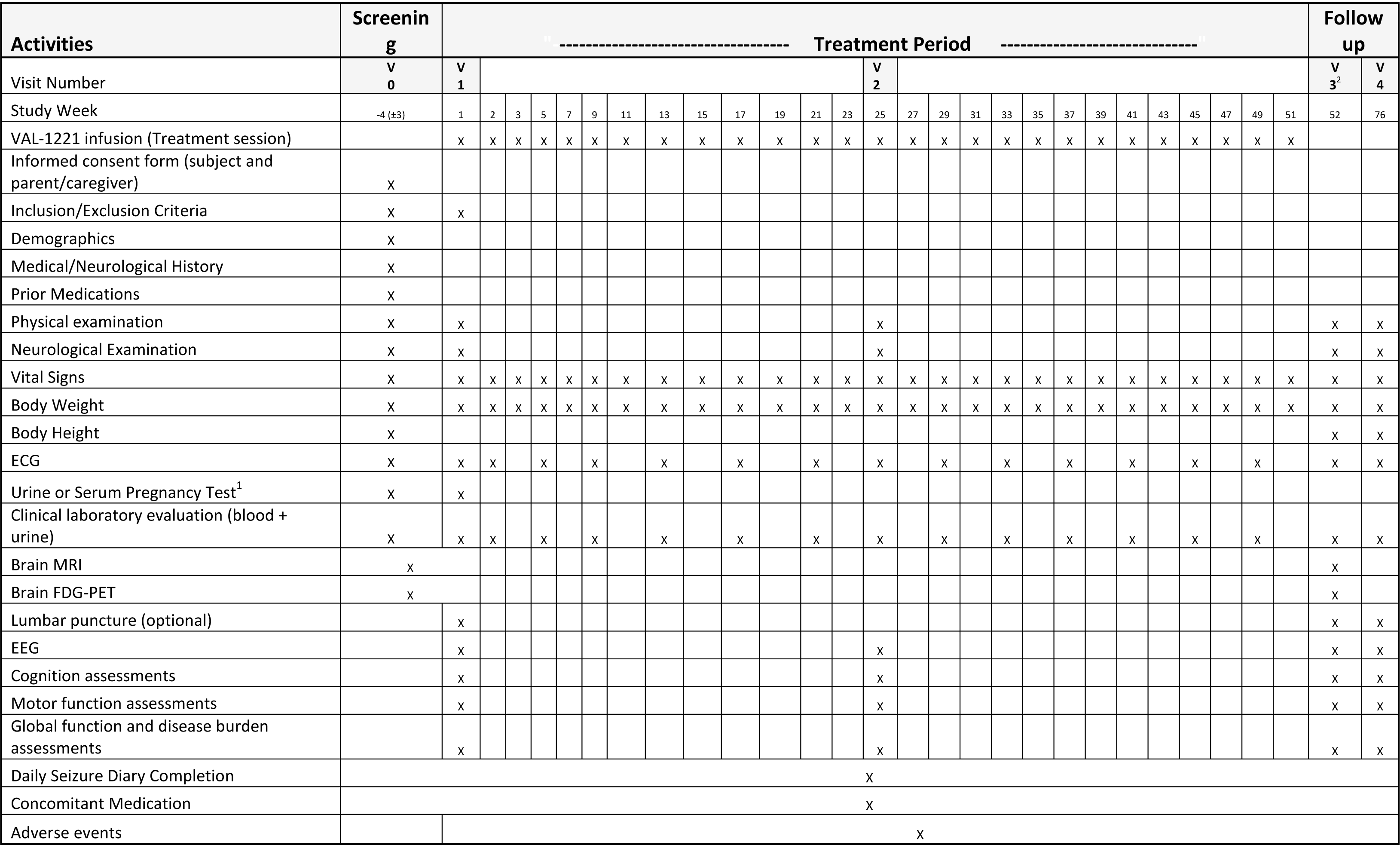
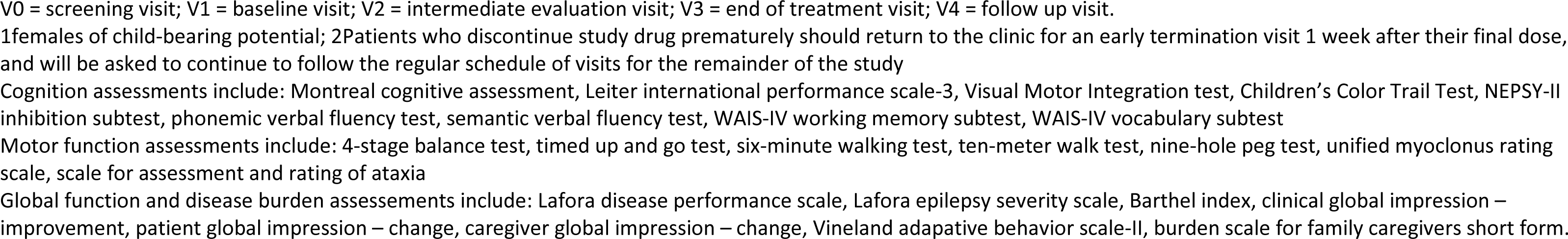
Schedule of study procedures.

Each patient will be required to undergo the following assessments during the screening, baseline, treatment, and follow-up phases at the study times indicated: vital signs measurement, 12-lead electrocardiogram (ECG), body weight and height, urine or serum pregnancy test (females only), laboratory evaluation (serum chemistry, hematology, urinalysis, etc.), physical and neurological examination, completion of seizure diary, concomitant medications recording, adverse event recording, EEG, brain MRI, brain FDG-PET, lumbar puncture (optional, only with additional consent provision), neuropsychological tests including Montreal Cognitive Assessment (MoCA), Leiter Performance Scale 3 (Leiter-3), Visual Motor Integration test (VMI), Children’s Color Trail Test (CCTT), NEPSY-II Inhibition subtest, phonemic and semantic verbal fluency test, WAIS-IV working memory and vocabulary subtests; motor functions tests including 4-stage balance test, timed up and go, 6-minute walk test, 10-meter walk test, 9-hole peg test, scale for assessment and rating of ataxia (SARA), unified myoclonus rating scale (UMRS); global and burden scales including Lafora Disease Performance Scale (LDPS), Lafora Epilepsy Severity Scale (LESS), Barthel Index, Vineland-II, burden scale for family caregivers (short form); change of clinical global impression scale-improvement (CGI-I), patient global impression scale-change (PGI-C), caregiver global impression scale-change (CaGI-C).

Every attempt should be made to have the same assessor administer the assessments to a given patient throughout the study.

### Early termination visit (data retention)

All patients who discontinue treatment or withdraw from the study early (during the open-label treatment period) will be asked to return the week after the final dose of study drug in order to complete the early termination visit, which consists of all the procedures performed at V3 (end-of-treatment visit). In addition, patients who discontinue treatment will be asked to return for collection of safety and efficacy data according to the schedule of activities until the end of the treatment period. These patients will also be asked to participate in the follow-up clinical and safety visits.

### Data management

Case report forms (CRFs) will be completed for each study subject. The data will be reported on paper CRFs consistently to source documents, and afterwards they will be transferred into an electronic database. It is the Investigator’s responsibility to ensure the accuracy, completeness, legibility, and timeliness of the data reported in the subject’s CRF.

### Safety endpoints

The safety of VAL-1221 (primary objective) will be evaluated on the basis of the following endpoints:

- Nature, incidence, seriousness, severity, and timing of AEs, including IRRs
- Incidence of treatment discontinuation due to adverse events
- Changes in vital signs, ECG and laboratory assessments from baseline over time and incidence of abnormalities

### Efficacy endpoints

The exploratory efficacy objective for VAL-1221 will be evaluated on the basis of the following endpoints, calculated from baseline to week 52 (12 months).

- Epilepsy: change in tonic-clonic seizures frequency comparing the trial period with the 6 months preceding the baseline, change in frequency of epileptiform discharges on EEG.
- Cognitive functions: non-worsening or improvement of MoCA, Leiter Performance Scale 3, Visual Motor Integration test, Children’s Color Trail Test, NEPSY-II Inhibition subtest, phonemic verbal fluency test, semantic verbal fluency test, WAIS-IV working memory subtest, WAIS-IV vocabulary subtest.
- Motor functions: non-worsening or improvement of 4-stage balance test, timed up and go, 6-minute walk test, 10-meter walk test, 9-hole peg test, SARA, UMRS
- Global assessment and disease burden: non-worsening or improvement of LDPS, LESS, Barthel Index, Vineland-II, Burden Scale for Family Caregivers (short form); change of CGI-I, PGI-C, Cagi-C.

### Biomarker endpoints

The aim is to evaluate biomarkers that can provide evidence of VAL-1221 activity or can increase the knowledge and understanding of disease biology and drug safety, such as: blood and CSF biomarkers (metabolic profile, markers of neurodegeneration and neuronal injury such as neurofilament light chain, total tau), brain MRI measures, brain FDG-PET measures.

### Statistical analyses

Statistical analyses will be primarily descriptive. Baseline data shall be summarized using the statistics n, median, mean, std, min, max, for all continuous variables and as n, percent (%) for each categorical variable. These descriptive summaries shall also be generated by visit for all endpoints. For the analysis of the continuous endpoints, summary statistics (n, mean, std, min, max 95% confidence interval [CI]) of the change from baseline to the specified time points shall be generated. The categorical endpoints shall be summarized using the proportion and 95% CI. Similar summary statistics for change from baseline to the specified time points shall be generated as appropriate.

All treated patients will be included in the analyses unless otherwise specified. If the patient has received any study treatment, all available safety data will be used

### Safety data analysis

Any unfavorable and unintended clinical finding, reported by the patient or revealed by any diagnostic procedure, including clinically relevant change in laboratory assessments and ECG, will be recorded as AE. This includes any newly occurring event or previous condition that has increased in severity or frequency since the administration of study drug (treatment-emergent).

The safety analysis will be performed on the basis of the above-mentioned safety endpoints. AEs will be summarized by nature, seriousness, severity, and relationship to the study drug, in addition to timing for IRRs. The frequency of AEs plus the number of patients experiencing an AE will be summarized.

### Efficacy and biomarkers data analysis

The exploratory efficacy and biomarker objectives include endpoints which represent key measures of disease progression to determine a potential treatment-related improvement.

To evaluate the reduction of seizure frequency, we will compare the frequency of tonic-clonic seizures as recorded during the 6 months preceding the baseline visit and the frequency in the 6 months preceding the end-of-treatment visit (week 52). Sub-analysis will be performed comparing the 6-month preceding the baseline visit to intermediate visit (week 25) and to follow-up visit (week 76). The halt or improvement in LD symptom progression (i.e., motor function, cognition, global assessment and disease burden) will be reported at the time points 25 weeks (interim analysis), 52 weeks and 76 weeks and compared with the baseline evaluation. Similarly, biomarker endpoints (blood and CSF analysis, brain MRI and FDG-PET) obtained at 52 weeks will be compared with baseline and categorized as improved, stable, and deteriorated.

When available, published results from the LD natural history cohort study (NCT03876522) regarding the change in outcomes in common with ours at baseline and at 52 weeks will be compared statistically.

### Data and safety monitoring board

A data and safety monitoring board of at least three independent qualified experts will be appointed before the start of the trial (neuropharmacologist, epileptologist, biostatistician).

### Interim analysis

An interim analysis at 25 weeks (approximately six month) will be performed. The key evaluation is safety and tolerability, but efficacy readouts will also be assessed.

### Protocol amendments

Protocol amendments that impact patient safety, change the scope of the investigation, or affect the scientific quality of the study must be approved by the ethics committee and submitted to the appropriate regulatory authorities before implementation of such modifications to the study. All investigators will receive a copy of the amended protocol.

In the event that the protocol needs to be modified immediately to eliminate an apparent hazard to a patient, the sponsor will amend and implement the protocol change and subsequently notify the regulatory authorities and/or the ethics committee, as appropriate.

### Patient confidentiality

To maintain patient privacy, all CRFs, banked study samples, study drug accountability records, study reports and communications will identify the patient by the assigned patient identification number. The patient’s confidentiality will be maintained and will not be made publicly available to the extent permitted by the applicable laws and regulations

## QUALITY CONTROL AND QUALITY ASSURANCE

Quality assurance and quality control systems with written standard operating procedures (SOPs) will be followed to ensure this trial will be conducted and data will be generated, documented (recorded), and reported in compliance with the protocol, Good Clinical Practice (GCP), and the applicable regulatory requirements.

The site’s dedicated study monitor will arrange to visit the investigator at regular intervals during the study. The monitoring visits will be conducted according to the applicable International Conference on Harmonization (ICH) and GCP guidelines to ensure protocol adherence, quality of data, drug accountability, compliance with regulatory requirements, and continued adequacy of the investigational site and its facilities.

During these visits, CRFs and other data related to the study will be reviewed and any discrepancies or omissions will be identified and resolved. The study monitor will be given access to study relevant source documents (including medical records) for purposes of source data verification.

During and/or after completion of the study the regulatory authorities may wish to perform on-site audits. The investigator will cooperate with any audit and provide assistance and documentation (including source data) as requested.

Quality control will be applied to each stage of data handling to ensure that all data are reliable and have been processed correctly.

The detailed monitoring activities, including data verification and revision, will be written in the Trial Monitoring Plan.

## ETHICS AND DISSEMINATION

The study will be conducted in accordance with standards of good clinical practice (GCP), as defined by The International Council for Harmonisation of Technical Requirements for Pharmaceuticals for Human Use (ICH), the principles of the declaration of Helsinki, and all applicable national and local regulations. The study protocol was approved by the local ethics committee (number 232-2023-FARM-AUSLBO–23020, 22-Mar-2023).

The principal investigator and the sponsor have no financial or competing interests for this trial and will have access to the final trial dataset without limitations.

The results of this study will be disseminated by the investigators through presentations at international scientific conferences and reported in peer-reviewed scientific journals, or in any case made publicly available (e.g., preprint repositories, communications to rare disease communities). Access to the full protocol and participant-level dataset will be granted upon reasonable request.

## WRITTEN INFORMED CONSENT

Written informed consent will be required from each patient prior to any testing under this protocol, including Screening tests and evaluations, according to the regulatory requirements and GCP. Minors aged 12 to 17 years will be administered an ad hoc ICF.

The background of the proposed study and the benefits and risks of the procedures and study will be explained to the patients. It is the responsibility of the investigator to obtain consent and to provide the patient with a copy of the signed and dated informed consent form (ICF). Confirmation of a patient’s informed consent must also be documented in the patient’s medical record prior to any testing under this protocol, including screening tests and evaluations.

For children/minors and subjects with intellectual disability unable to give informed consent, the ICF will be administered to the parents or other legal representatives.

All ICFs used in this study was approved by the appropriate IEC.

## CONFIDENTIALITY

To maintain patient privacy, all CRFs, banked study samples, study drug accountability records, study reports and communications will identify the patient by the assigned patient identification number. The investigator will grant monitor(s) and auditor(s) from the sponsor or designee and regulatory authority(ies) access to the patient’s original medical records for verification of data gathered on the CRFs and to audit the data collection process. The patient’s confidentiality will be maintained and will not be made publicly available to the extent permitted by the applicable laws and regulations

## DISCUSSION

Lafora disease is severe, progressive, and ultimately fatal in late teenage years or by early adulthood. The potential benefit of VAL-1221, the demonstrated lack of serious or unexpected AEs with VAL-1221 in clinical studies, the seriousness of LD and the lack of alternative treatment options, warrants a trial of VAL-1221 in patients with LD.

This study aims to determine the safety and preliminary efficacy of VAL-1221 administered intravenously in patients with LD. This was the first approved study protocol developed for a potential disease-modifying drug in LD.

VAL-1221 is able to clear systemic and brain LBs after intravenous and intracerebroventricular administration, respectively (under publication, personal communication of the corresponding Author Prof. Matthew Gentry, University of Florida). Although the current VAL-1221 formulation is not suitable for human ICV injections, intravenous VAL-1221 could reasonably reach the brain in quantities sufficient for a meaningful response via ENT2, facilitating brain endothelial cell penetration and BBB transport (Rattray et al 2021), and BBB disruption due to tonic-clonic seizures (Cudna et al., 2023), which are frequent in LD.

The rationale for VAL-1221 dose and schedule comes from experience in Pompe disease clinical trials (Kishnani et al., 2009) (Kishnani et al., 2019) and expanded access programs, which are using the same treatment scheme.

The study will be open label with a 12-month treatment phase. The decision not to include a placebo control group was taken due to the serious and life-threatening nature of the disease and its rarity. This happens quite commonly in diseases with similar features without alternative treatment options as, if a new therapy is promising, it might be unethical to have patients get a placebo (May, 2023). Given the lack of a control group in this study, the exploratory endpoints observed will be baseline-controlled, i.e., will be compared during and after the trial to a baseline determined at the start of the trial. As LD is relentlessly progressive, any arrest or improvement of patients’ clinical deficits should be conspicuous and would not be plausibly explained by factors other than treatment with VAL-1221.

Another possibility to determine efficacy for single-arm trials is to use a synthetic control arm, wherein a placebo group is modeled on the basis of previously collected real-world data (Pizzamiglio et al., 2022). The results of an international natural history study of LD (NCT03876522), which has followed-up 33 patients over 24 months, should be published soon. Thus, we decided to include in our study protocol standardized biomarkers and clinical outcome measures already used in the natural history study (including EEG, LDPS, Leiter Performance Scale 3, Children’s Color Trail Test, timed up and go, 6-minute walk test, 9-hole peg test, SARA, Vineland-II, etc.), in order to obtain meaningful insight of the effects of VAL-1221 by comparing the two groups, trying to select those with similar disease characteristics, including severity, illness duration and prior treatments. The application of natural history data for drug development is not limited to the provision of an external control population, but may also be pivotal in the development and validation of biomarkers and clinical outcome measures (Pizzamiglio et al., 2022).

Additionally, we decided to include additional outcomes measures not assessed in the natural history study that might be helpful in providing evidence of VAL-1221 activity. These include brain FDG-PET, which has been shown to be highly sensitive to evaluate LD at any stage and may correlate with disease progression (Muccioli et al., 2020, d’Orsi et al., 2023), as well as neuropsychological tests assessing frontal lobe-related functions, such as executive functions, which are predominantly involved in LD (Pichiecchio et al. 2008).

Based upon VAL-1221 compassionate treatment data, and preliminary analysis of the natural history study (*unpublished*), a 12-month treatment duration was considered sufficient to assess the safety and exploratory efficacy objectives.

The total sample size of 6 patients is not based on a formal statistical calculation; it is a relatively small sample size, partially offset by the rarity of the condition, which is related to the availability of the drug product VAL-1221. If additional funding becomes available, it would be possible to enroll a larger number of patients with LD. This would allow for more robust statistical analysis and a better understanding of the potential benefits of VAL-1221.

In summary, this study protocol for VAL-1221 in LD represents a crucial first step towards developing effective treatments for this devastating condition. The results of this study could inform the preparation of future study protocols and ultimately contribute to improving the lives of LD patients.

## Trial Status

The trials is registered on the European Union Clinical Trials Register with EudraCT number 2023-000185-34. The trials is not yet open for participant recruitment.

## Data Availability

All data produced in the present work are contained in the manuscript

## Acknowledgements

Prof. Emilio Perucca for his precious advices and Eleonora Romagnoli for her assistance in obtaining protocol approvals. A.I.LA. Associazione Italiana Lafora OdV and Associazione Malattie Rare Mauro Baschirotto for their unconditional support.

## Author contributions

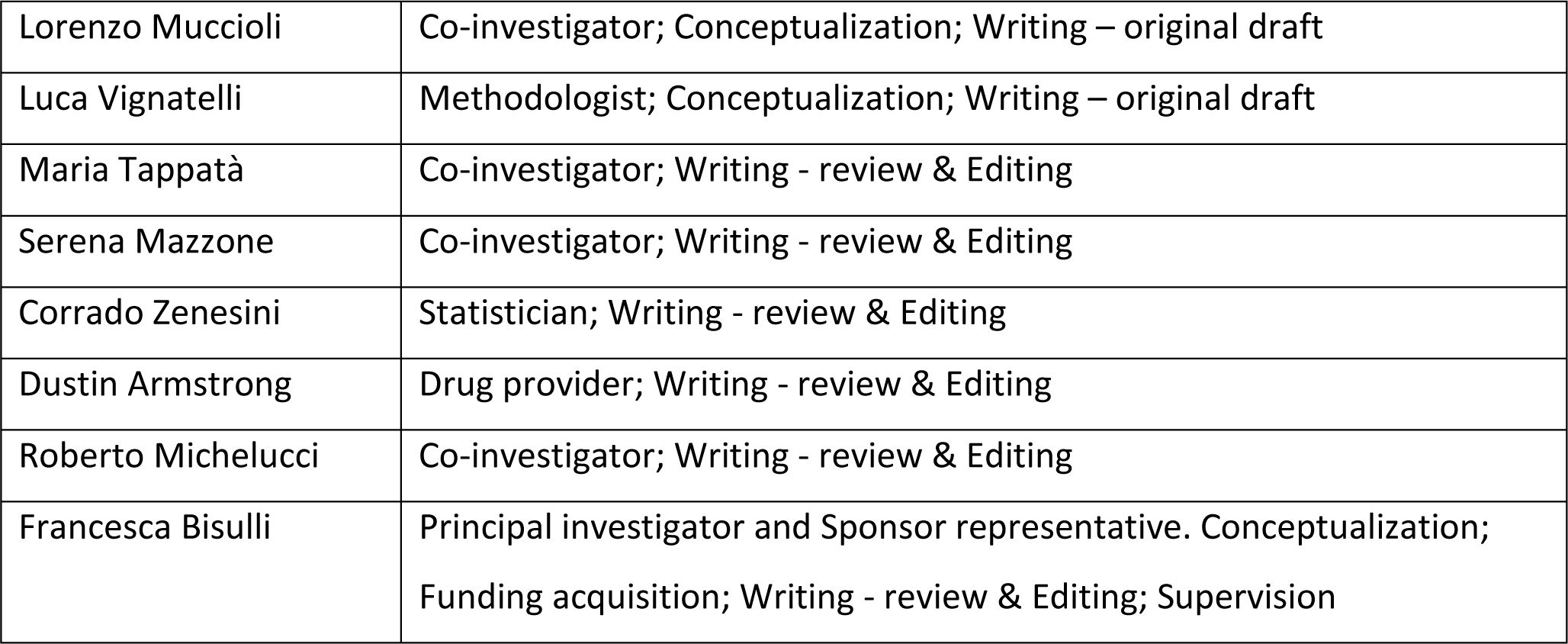

## Collaborators of the DEFEAT-LD Study Group

Cosimo Altomare^1^; Massimo Carella^2^; Cinzia Costa^3,4^; Giuseppe Damante^5,6^; Lidia Di Vito^7^; Giuseppe d’Orsi^2^; Nicola Gambacorta^2^; Paola Imbrici^1^; Valentina Imperatore^3,8^; Antonella Liantonio^1^; Laura Licchetta^7^; Raffaele Lodi^7^; Paola Mantuano^1^; Orazio Palumbo^2^; Elena Pasini^7^; Paolo Prontera^8^

1. Dipartimento di Farmacia-Scienze del Farmaco, Università degli studi di Bari Aldo Moro, Bari, Italy
2. Fondazione IRCCS-Casa Sollievo della Sofferenza, San Giovanni Rotondo, Italy
3. Department of Medicine and Surgery, University of Perugia, Perugia, Italy
4. Section of Neurology, S. Maria della Misericordia Hospital, Perugia, Italy
5. Department of Medicine (DAME), University of Udine, Udine, Italy
6. Institute of Medical Genetics, Udine University Hospital, Udine, Italy
7. IRCCS Istituto delle Scienze Neurologiche di Bologna, Full Member of the European Reference Network for Rare and Complex Epilepsies (EpiCARE), Bologna, Italy
8. Medical Genetics Unit, S. Maria della Misericordia Hospital, Perugia, Italy.

## Data statement

All information concerning this clinical trial is property of the Sponsor and the investigators. The results will be published in peer-reviewed journals, or in any case made publicly available (e.g. preprint repositories).

## REFERENCES

Chan AW, Tetzlaff JM, Altman DG, et al. SPIRIT 2013 statement: defining standard protocol items for clinical trials. Ann Intern Med. 2013;158(3):200-207. doi:10.7326/0003-4819-158-3-201302050-00583

Cudna A, Bronisz E, Jopowicz A, Kurkowska-Jastrzębska I. Changes in serum blood-brain barrier markers after bilateral tonic-clonic seizures. Seizure. 2023;106:129–137. doi:10.1016/j.seizure.2023.02.012

DePaoli-Roach AA, Tagliabracci VS, Segvich DM, Meyer CM, Irimia JM, Roach PJ. Genetic depletion of the malin E3 ubiquitin ligase in mice leads to lafora bodies and the accumulation of insoluble laforin. J Biol Chem. 2010;285(33):25372–25381. doi:10.1074/jbc.M110.148668

d’Orsi G, Farolfi A, Muccioli L, et al. Association of CSF and PET markers of neurodegeneration with electroclinical progression in Lafora disease. Front Neurol. 2023;14:1202971. Published 2023 Jun 28. doi:10.3389/fneur.2023.1202971

Duran J, Gruart A, García-Rocha M, Delgado-García JM, Guinovart JJ. Glycogen accumulation underlies neurodegeneration and autophagy impairment in Lafora disease. Hum Mol Genet. 2014;23(12):3147–3156. doi:10.1093/hmg/ddu024

Franceschetti S, Gambardella A, Canafoglia L, et al. Clinical and genetic findings in 26 Italian patients with Lafora disease. Epilepsia. 2006;47(3):640–643. doi:10.1111/j.1528-1167.2006.00479.x

Ganesh S, Delgado-Escueta AV, Sakamoto T, et al. Targeted disruption of the Epm2a gene causes formation of Lafora inclusion bodies, neurodegeneration, ataxia, myoclonus epilepsy and impaired behavioral response in mice. Hum Mol Genet. 2002;11(11):1251–1262. doi:10.1093/hmg/11.11.1251

Kishnani PS, Corzo D, Leslie ND, et al. Early treatment with alglucosidase alpha prolongs long-term survival of infants with Pompe disease. Pediatr Res. 2009;66(3):329–335. doi:10.1203/PDR.0b013e3181b24e94

Kishnani P, Lachmann R, Mozaffar T, et al. Safety and efficacy of VAL-1221, a novel fusion protein targeting cytoplasmic glycogen, in patients with late-onset Pompe disease. Molecular Genetics and Metabolism. 2019;126(2), S85-S86.

Llerena Junior JC, Nascimento OJ, Oliveira AS, et al. Guidelines for the diagnosis, treatment and clinical monitoring of patients with juvenile and adult Pompe disease [published correction appears in Arq Neuropsiquiatr. 2016 Feb;74(2):VI]. Arq Neuropsiquiatr. 2016;74(2):166-176. doi:10.1590/0004-282X20150194

May, M. Rare-disease researchers pioneer a unique approach to clinical trials. Nat Med 2023;29 1884–1886. doi: 10.1038/s41591-023-02333-4

Muccioli L, Farolfi A, Pondrelli F, et al. FDG-PET assessment and metabolic patterns in Lafora disease. Eur J Nucl Med Mol Imaging. 2020;47(6):1576–1584. doi:10.1007/s00259-019-04647-3

Nitschke F, Ahonen SJ, Nitschke S, Mitra S, Minassian BA. Lafora disease - from pathogenesis to treatment strategies. Nat Rev Neurol. 2018;14(10):606–617. doi:10.1038/s41582-018-0057-0

Pichiecchio A, Veggiotti P, Cardinali S, Longaretti F, Poloni GU, Uggetti C. Lafora disease: spectroscopy study correlated with neuropsychological findings. Eur J Paediatr Neurol. 2008;12(4):342–347. doi:10.1016/j.ejpn.2007.09.008

Pizzamiglio C, Vernon HJ, Hanna MG, Pitceathly RDS. Designing clinical trials for rare diseases: unique challenges and opportunities. Nat Rev Methods Primers. 2022;2(1):s43586–022-00100-2. Published 2022 Mar 10. doi:10.1038/s43586-022-00100-2

Pondrelli F, Muccioli L, Licchetta L, et al. Natural history of Lafora disease: a prognostic systematic review and individual participant data meta-analysis. Orphanet J Rare Dis. 2021;16(1):362. Published 2021 Aug 16. doi:10.1186/s13023-021-01989-w

Rattray Z, Deng G, Zhang S, et al. ENT2 facilitates brain endothelial cell penetration and blood-brain barrier transport by a tumor-targeting anti-DNA autoantibody. JCI Insight. 2021;6(14):e145875. Published 2021 Jul 22. doi:10.1172/jci.insight.145875

Turnbull J, Tiberia E, Striano P, et al. Lafora disease. Epileptic Disord. 2016;18(S2):38-62. doi:10.1684/epd.2016.0842

Zhou Z, Austin GL, Shaffer R, Armstrong DD, Gentry MS. Antibody-Mediated Enzyme Therapeutics and Applications in Glycogen Storage Diseases. Trends Mol Med. 2019;25(12):1094–1109. doi:10.1016/j.molmed.2019.08.005

